# Development and validation of imaging-free myocardial fibrosis prediction models, association with outcomes, and sample size estimation for phase 3 trials

**DOI:** 10.1101/2024.02.07.24302443

**Authors:** Nicholas Black, Joshua Bradley, Gavin Lewis, Jakub Lagan, Christopher Orsborne, Fardad Soltani, John P. Farrant, Theresa McDonagh, Matthias Schmitt, João L Cavalcante, Martin Ugander, Javed Butler, Mark C. Petrie, Christopher A. Miller, Erik B. Schelbert

## Abstract

**Background and Aims:** Phase 3 trials testing whether pharmacologic interventions targeting myocardial fibrosis (MF) improve outcomes require MF measurement that does not rely on tomographic imaging with intravenous contrast.

**Methods:** We developed and externally validated extracellular volume (ECV) prediction models incorporating readily available data (comorbidity and natriuretic peptide variables), excluding tomographic imaging variables. Survival analysis tested associations between predicted ECV and incident outcomes (death or hospitalization for heart failure). We created various sample size estimates for a hypothetical therapeutic clinical trial testing an anti-fibrotic therapy using: a) predicted ECV, b) measured ECV, or c) no ECV.

**Results:** Multivariable models predicting ECV had reasonable discrimination (optimism corrected C-statistic for predicted ECV ≥27% 0.78 (95%CI 90.75-0.80) in the derivation cohort (n=1663) and 0.74 (95%CI 0.71-0.76) in the validation cohort (n=1578)) and reasonable calibration. Predicted ECV associated with adverse outcomes in Cox regression models: ECV ≥27% (binary variable) HR 2.21 (1.84–2.66). For a hypothetical clinical trial with an inclusion criterion of ECV ≥27%, use of predicted ECV (with probability threshold of 0.69 and 80% specificity) compared to measured ECV would obviate the need to perform 3940 CMR scans, at the cost of an additional 3052 participants screened and 705 participants enrolled.

**Conclusions:** Predicted ECV (derived without tomographic imaging) associates with outcomes and efficiently identifies vulnerable patients who might benefit from treatment. Predicted ECV may foster the design of phase 3 trials targeting MF with higher numbers of screened and enrolled participants, but with simplified eligibility criteria, avoiding the complexity of tomographic imaging.

**Structured Graphical Abstract:** *Key Question:* Phase 3 trials targeting myocardial fibrosis (MF) to improve outcomes require MF measurement that does not rely on tomographic imaging with intravenous contrast. So, we developed and validated extracellular volume (ECV) prediction models incorporating clinical data, excluding tomographic imaging.

*Key Finding:* Predicted ECV had reasonable discrimination and associated with outcomes. For a hypothetical trial with an ECV ≥27% inclusion criterion, using predicted ECV versus measured ECV would avoid 3940 cardiovascular magnetic resonance (CMR) scans, but require an additional 3052 participants screened and 705 enrolled.

*Take-home Message:* Predicted ECV (derived without imaging) associates with outcomes and efficiently identifies vulnerable patients. Predicted ECV may foster phase 3 trials targeting MF with higher numbers of screened and enrolled participants, but simplified eligibility criteria, avoiding the complexity of tomographic imaging. 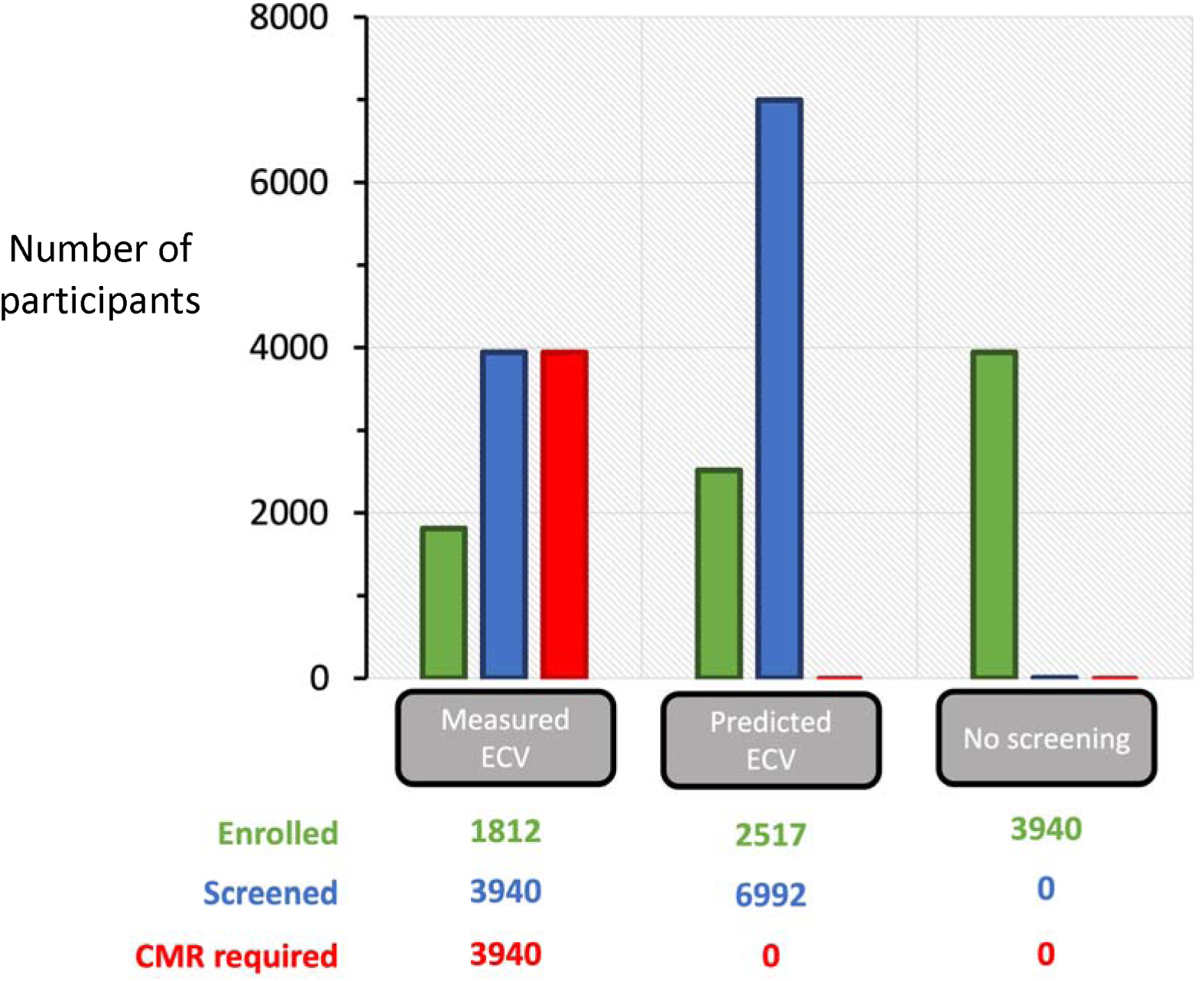
For a hypothetical trial requiring 1812 participants with measured ECV ≥27%, 3940 patients would need to undergo screening with CMR. If predicted ECV is used, an additional 3052 patients would need to be screened and an additional 705 patients enrolled, but no patients would require CMR. If no screening is used, an additional 2128 patients would need to be enrolled.

## Introduction

Phase 3 trials testing whether pharmacologic interventions targeting myocardial fibrosis (MF) improve outcomes require MF measurement that does not rely on tomographic imaging. MF is prevalent across many different disease states (e.g., heart failure regardless of systolic function, hypertension, diabetes, valvular disease, etc.) but not universal, so historically tomographic imaging has been required to identify those with MF. Over past decades, abundant mechanistic, preclinical and clinical studies implicate MF as not simply a marker of risk, but rather a causal disease process originating from fibroblast dysregulation, culminating in myocardial remodelling and ultimately morbidity and mortality. As such, multiple pharmacologic agents targeting MF are in development.

Extracellular volume (ECV) measurement represents the best validated measure of MF (1,2) that robustly associates with outcomes.(3-5) Yet, ECV requires tomographic imaging such as cardiovascular magnetic resonance (CMR) or computed tomography (CCT). These imaging modalities incur significant expense and expose patients to intravenous contrast agents. As such tomographic imaging is suitable for phase 2, but not phase 3 clinical trials. It is implausible that phase 3 trials of anti-fibrotic therapies would enrol patients based on costly and complex imaging techniques that could limit eligibility criteria. Rather phase 3 trials require simplified strategies for estimating MF based on multivariable inputs of ubiquitous and inexpensive data elements, such as comorbidity and laboratory variables.

To foster the eventual implementation of phase 3 trials targeting MF without imaging, we leveraged large, prospectively assembled CMR cohorts that measured ECV routinely. We developed and externally validated ECV prediction models incorporating readily available data elements, excluding tomographic imaging variables. We hypothesized that *predicted* ECV would associate with incident hospitalisation for heart failure or all-cause mortality. Finally, to demonstrate how predicted ECV could impact phase 3 trial design, we created a variety of sample size estimates for a hypothetical therapeutic clinical trial testing an anti-fibrotic therapy using: a) predicted ECV, b) measured ECV, or c) no ECV.

## Methods

### Study population

Between June 1, 2016, and May 31, 2018, consecutive adult patients undergoing clinically indicated CMR imaging at Manchester University National Health Service Foundation Trust (MFT), Manchester, UK, were prospectively recruited into a development cohort (registered at <u>ClinicalTrials.gov</u>, <u>NCT02326324</u>). Exclusion criteria included a diagnosis of any of the following: amyloidosis, complex congenital heart disease, Fabry disease, hypertrophic cardiomyopathy, iron overload, myocarditis, and stress-induced cardiomyopathy. Patients were also excluded if their CMR scan was not suitable for analysis. The investigation conforms with the *Declaration of Helsinki.* The study was approved by a NHS Research Ethics Committee, and all participants provided written informed consent.

### Procedures

Data were managed by use of Research Electronic Data Capture (known as REDCap).(6) Baseline characteristics were identified from medical records. CMR was done on a single 1.5T scanner (1·5 T Avanto; Siemens Medical Imaging; Erlangen, Germany). Scanning comprised steady-state free precession cine imaging in standard long axis and short axis planes, basal and mid left ventricle short axis T1 mapping (MOdified Look-Locker Inversion Recovery) imaging before and after administration of a gadolinium-based contrast agent (gadoterate meglumine [Dotarem]; Guerbet; Paris, France), and late gadolinium enhancement imaging. CMR analysis was done by use of <u>cvi42</u> (version 5.6.7; Circle Cardiovascular Imaging; Calgary, AB, Canada). Measurements of ventricular mass, volumetrics, ejection fraction, and atrial area were done in accordance with recommendations from the Society for Cardiovascular Magnetic Resonance.(7) Global longitudinal strain was measured as described previously.(8) Myocardial fibrosis was measured by use of the extracellular volume (ECV) technique, according to recommendations from the Society for Cardiovascular Magnetic Resonance.(9) N-terminal pro-B-type natriuretic peptide (NT-pro-BNP) was laboratory assessed from blood sampling performed on the same day as CMR (cobas e 411 immunoanalyser; Roche Diagnostics; Mannheim, Germany). All baseline data collection, including the CMR analysis, was done before receiving, and was therefore blinded to, the outcome data.

### Model development

Model development is reported in accordance with Transparent Reporting of a Multivariable Prediction Model for Individual Prognosis or Diagnosis guidelines.(10) Candidate predictor variables were selected according to clinical practice and literature review. CMR variables were excluded from model development as the purpose was to predict ECV without the use of CMR. In total, 17 candidate predictor variables were considered: age, female sex, white race, body mass index (BMI), myocardial infarction (MI), stroke, diabetes mellitus (DM), hypertension (HTN), hyperlipidaemia, chronic obstructive pulmonary disease (COPD), atrial fibrillation (AF), current smoker, ex-smoker, prior heart failure (HF) hospitalisation, estimated glomerular filtration rate (eGFR), N-terminal pro B-type natriuretic peptide (NT-proBNP) and QRS duration. Natural logarithm (*ln*) transformation of NT-proBNP was performed in keeping with previous studies.(11)

We developed our model in a development cohort of 1663 patients. Missing data for the candidate predictor variables was rare (4.1%), although 47% of patients were missing at least one value (Supplementary Table 1). Missing data were unintentional and their absence was due to incomplete medical records, incomplete CMR, or blood sampling not being done; thus, data were assumed to be missing at random. Multiple imputation by chained equations was used to create 47 imputed datasets.(12) Missing data were imputed from the candidate predictor and outcome variables by use of predictive mean matching.

Two models for predicting ECV were derived: i) a multivariable linear regression model for predicting ECV as a continuous variable ii) a multivariable logistic regression for predicting ECV ≥27% (binary variable). ECV ≥27% was the eligibility criterion for PIROUETTE, a phase 2 randomised trial which demonstrated a significant reduction in myocardial fibrosis in patients with heart failure with preserved ejection fraction treated with the anti-fibrotic drug pirfenidone.(13) ECV ≥27% equated to one standard deviation above the mean in healthy volunteers at the host institution (Manchester University NHS Foundation Trust). Assumptions for both models were satisfactory (Supplementary Figures 1 and 2). Stepwise model selection according to Akaike’s Information Criterion (AIC) was done separately in each imputed dataset. Variables present in more than 50% of the models were considered for inclusion in the parsimonious model by means of the Wald statistic test with P >0.1 as selection criterion.

### Internal validation

The developed model was validated internally. Non-parametric bootstrapping was used to estimate optimism and examine model stability. In each of the 47 imputed datasets, the entire modelling process, including predictor variable selection, was repeated in 1000 bootstrap samples. Model performance measures were pooled according to Rubin’s rule.(14) For the multivariable linear regression model, optimism-corrected R^2^, calibration slope and intercept, as well as mean absolute error (MAE) and root mean square error (RMSE), were calculated. For the multivariable logistic regression model, optimism-corrected Harrell’s C-statistic, calibration slope and calibration intercept were calculated. Calibration plots were inspected visually. Following internal validation, the optimism corrected calibration slope was used as a uniform shrinkage factor to reduce overfitting prior to external validation.(15)

### External validation

Between June 1, 2010, and March 25, 2016, consecutive adult patients undergoing clinically indicated CMR at the UPMC Cardiovascular Magnetic Resonance Center, in Pittsburgh, PA, USA, were prospectively recruited into a validation cohort. Exclusion criteria were identical to the derivation cohort. The study was approved by University of Pittsburgh Institutional Review Board, and all participants provided written informed consent. Missing data for the candidate predictor variables was rare (4.7%), although 87% of patients were missing at least one value (Supplementary Table 1). As for the derivation cohort, data was assumed to be missing at random. Multiple imputation by chained equations was used to create 87 imputed datasets. BNP (but not NT-pro-BNP) was available in the validation cohort, and given that they provide similar risk stratification, BNP was substituted for NT-pro-BNP in the external validation model.

In each of the 87 imputed datasets, the optimism corrected models were externally validated and model performance measures were pooled according to Rubin’s rule.(14). For the multivariable linear regression model, R^2^, calibration slope, calibration intercept, MAE and RMSE were calculated. For the multivariable logistic regression model, Harrell’s C statistic, calibration slope and calibration intercept were calculated. Calibration plots were inspected visually. The model was then recalibrated to the validation cohort by adjusting for the calibration slope and intercept from the external validation.(15)

### Prognostic modelling

Prognostic modelling was performed on the combined derivation (MFT) and validation (UPMC) cohorts. The outcome for prognostic modelling was a composite of hospitalisation for heart failure or all-cause mortality occurring after CMR. Time to the composite outcome was defined as time to first event, with censoring at the last follow-up date if no event occurred. Outcome data were obtained as described previously.(16) For the multivariable logistic regression model, a probability threshold of 0.69 was used to categorise patients as predicted ECV ≥27% with 80% specificity and 53% sensitivity (Supplementary Figure 7). The relationship between predicted ECV (either as a continuous variable or as a categorical variable ECV ≥ 27%) and the composite outcome was assessed by visual inspection of Kaplan-Meier curves and univariable analysis using Cox proportional hazards model.

### Sample size estimates

The multivariable logistic regression model could be used as the basis to recruit participants to a hypothetical therapeutic clinical trial with ECV ≥27% as an inclusion criterion. The following trial assumptions were made mirroring a recent HF trial(17): i) Superiority trial to determine the therapeutic benefit of a drug in patients with ECV ≥27%, ii) Single treatment and control group, iii) Type 1 error rate of 5%, iv) Statistical power of 80%, iv) Primary outcome occurs in 20% of the control group and 15% in the treatment group, v) Minimum clinical benefit of 5% absolute risk reduction or 25% relative risk reduction in the primary outcome, vi) Prevalence of observed ECV ≥27% of 46% (same as in combined derivation and validation cohorts).

Prior to enrolment into the hypothetical trial, sample size estimates for three different screening strategies are presented i) Screen participants with multivariable logistic regression model to identify participants with predicted ECV ≥27%, ii) Screen participants with CMR to identify participants with actual measured ECV ≥27%, iii) Enrolment without prior screening.

## Results

### Patient characteristics

The baseline characteristics of the derivation and validation cohorts are summarised in Table 1. Both measured and predicted ECV ≥27% were prevalent, 49.7% (1611/3241) and 47.6% (1543/3241), (McNemar’s test χ^2^ 4.4, p=0.036). The median follow-up duration was 1134 days (IQR 972 – 1353) in the derivation cohort and 2050 days (IQR 1486 – 2404) in the validation cohort. No patients were lost to follow-up or withdrew from the study. The mean myocardial ECV in the derivation and validation cohorts was 26.8% and 28.1%, respectively.

**Table 1:**
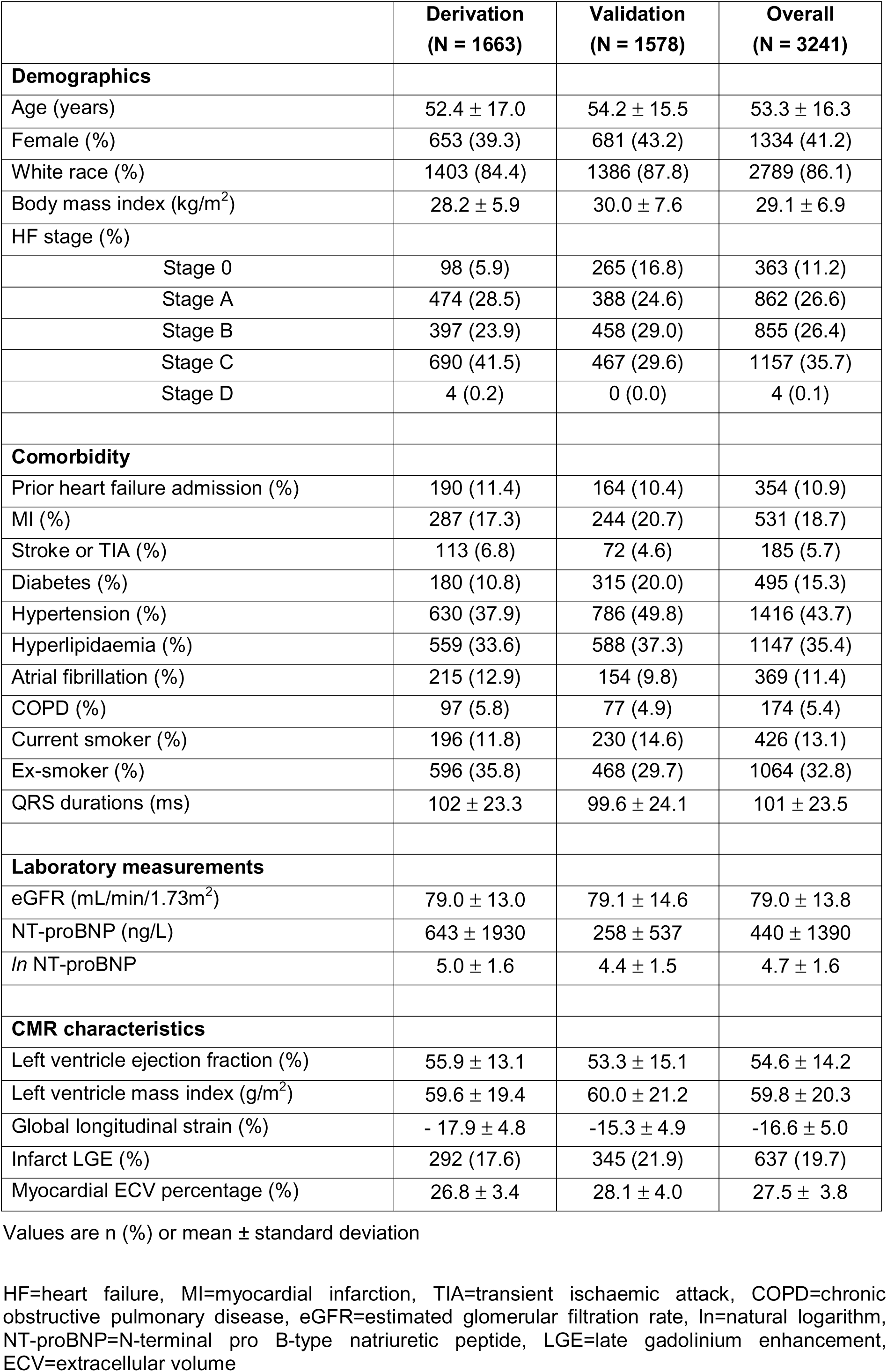
Table of baseline characteristics.

### Determinants of ECV as a continuous variable

Univariable associations between candidate predictor variables and myocardial ECV are presented in Supplementary Table 2. Predicted ECV (continuous variable) had a stronger association with measured ECV compared to other clinical covariates (Supplementary Table 2). Independent determinants of myocardial ECV were BMI, current smoking, eGFR, female sex, prior HF hospitalisation, *ln* NT-proBNP, white race, stroke and diabetes (Table 2 and Supplementary Table 3). *ln* NT-proBNP was the strongest predictor of measured ECV in multivariable models (Table 2). Metrics of model fit (R^2^, RMSE, MAE) and model calibration (intercept and slope) are shown in Supplementary Table 4. The optimism corrected R^2^ and 95% confidence interval (95% CI) for the model was 0.29 (0.26-0.33) in the derivation cohort and 0.24 (0.20-0.28) in the validation cohort. The corresponding calibration plots of observed vs. predicted ECV are shown in Supplementary Figures 3 and 4. The final model coefficients and calibration plot for the recalibrated model are shown in Table 2 and Figure 1, respectively.

**Figure 1:**
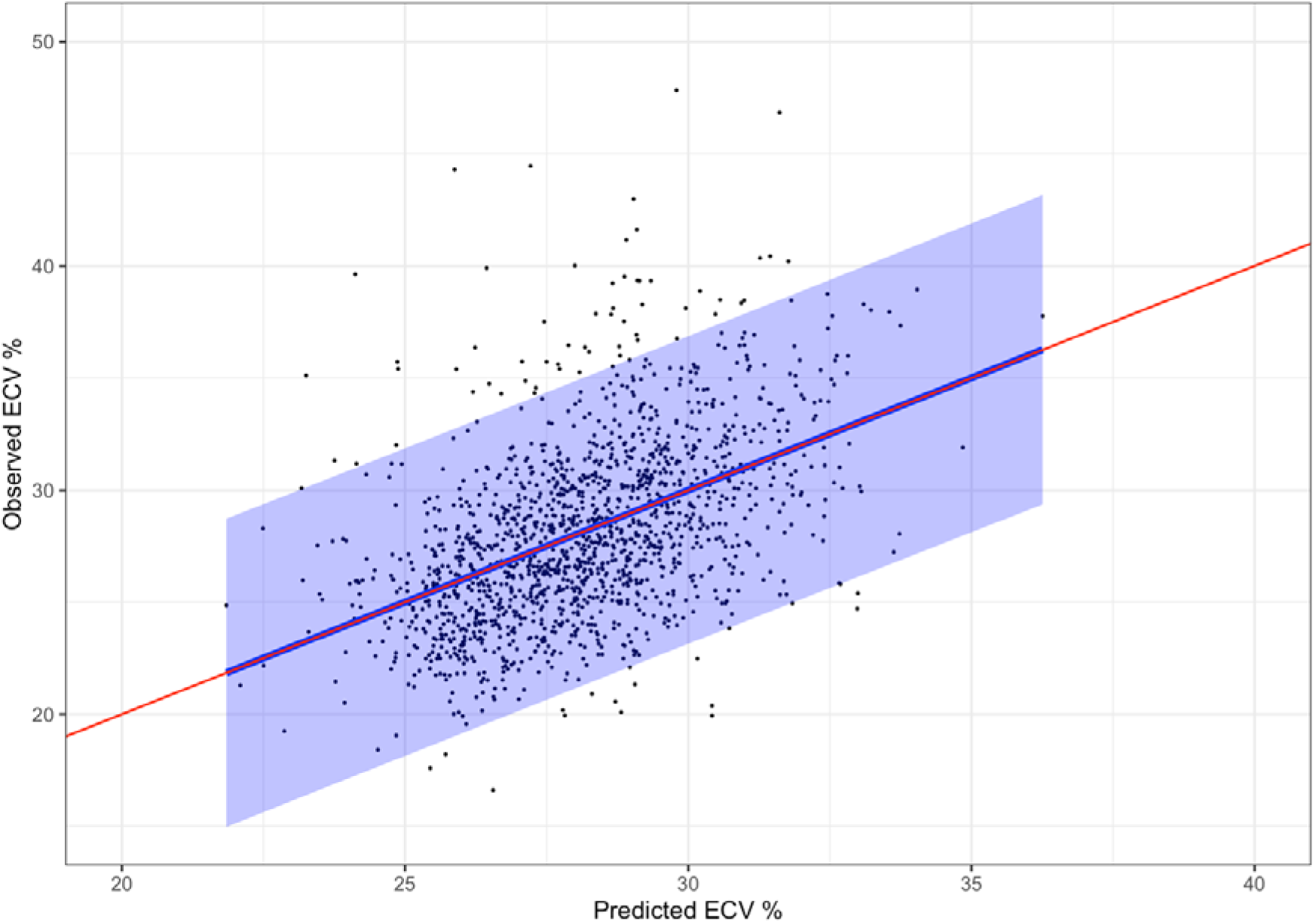
Calibration plot of recalibrated multivariable linear model for predicting ECV in validation cohort. Plot from last imputed dataset shown. Red is line of perfect prediction; blue is regression line and 95% prediction interval.

**Table 2:** Final recalibrated multivariable linear regression model for predicting ECV as a continuous variable.

| Variable | Recalibrated regression coefficients | 95% CI | T statistic | P value |
| --- | --- | --- | --- | --- |
| (Intercept) | 25.09 | 23.47 – 26.70 | 29.84 | <0.001 |
| BMI | -0.12 | -0.14 – -0.09 | -8.40 | <0.001 |
| Current smoker | 0.82 | 0.29 – 1.34 | 3.10 | <0.01 |
| eGFR | 0.03 | 0.01 – 0.04 | 3.97 | <0.001 |
| Female | 1.90 | 1.57 – 2.22 | 11.55 | <0.001 |
| Prior HF hospitalisation | 0.63 | 0.09 – 1.17 | 2.30 | 0.02 |
| *ln* NT-proBNP | 0.91 | 0.80 – 1.03 | 15.33 | <0.001 |
| White race | -0.77 | -1.22 – -0.32 | -3.42 | <0.001 |
| Stroke | -0.57 | -1.24 – 0.09 | -1.70 | 0.09 |
| DM | 0.60 | 0.06 – 1.15 | 2.20 | 0.03 |

ECV=extracellular volume, BMI=body mass index, CI=confidence interval, DM= diabetes mellitus, HF=heart failure, eGFR=estimated glomerular filtration rate, *ln*=natural logarithm, NT-proBNP=N-terminal pro B-type natriuretic peptide

### Determinants of ECV **≥** 27%

Univariable associations between candidate predictor variables and myocardial ECV ≥ 27% are presented in Supplementary Table 5. Predicted ECV (continuous variable) and predicted ECV ≥ 27% (binary variable) had a stronger association with measured ECV compared to other clinical covariates (Supplementary Table 5). Independent determinants of myocardial ECV ≥ 27% were BMI, eGFR, female sex, *ln* NT-proBNP, stroke, white race, COPD, prior HF hospitalisation and hypertension (Table 3 and Supplementary Table 6). Again, *ln* NT-proBNP, was the strongest predictor in multivariable models (Table 3). Metrics of model calibration (C-statistic, intercept, slope) are shown in Supplementary Table 7. The optimism corrected C-statistic and 95% CI for the model was 0.78 (0.75-0.80) in the derivation cohort and 0.74 (0.71-0.76) in the validation cohort. The corresponding calibration plots of observed vs. predicted ECV are shown in Supplementary Figures 5 and 6. The final model coefficients and calibration plot for the recalibrated model are shown in Table 3 and Figure 2, respectively.

**Figure 2:**
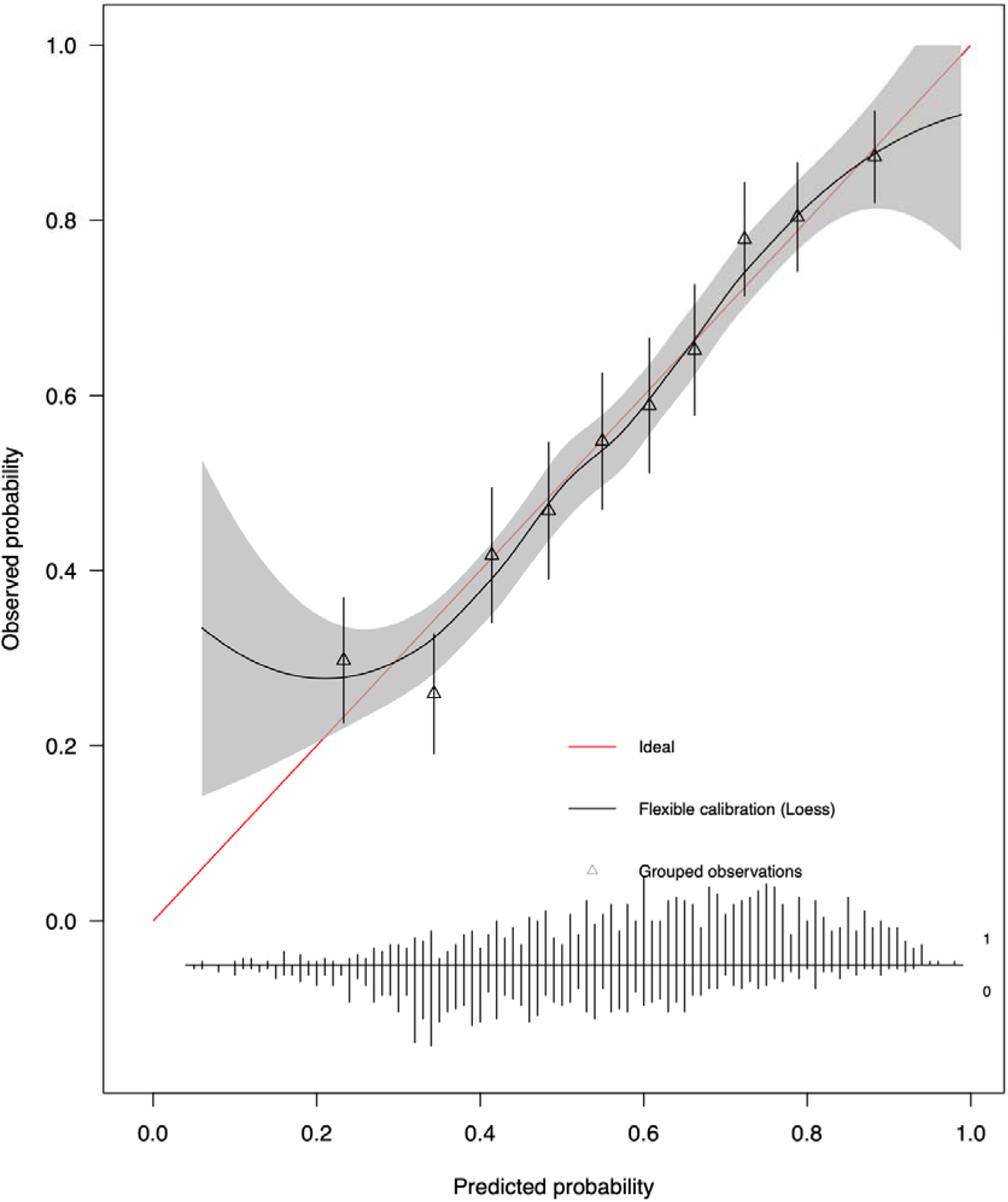
Calibration plot of recalibrated multivariable logistic regression model for predicting ECV ≥27% in validation cohort. Decile groups (triangle), smoothed Loess curve (grey shade) and line of perfect prediction (red line) illustrated. Plot from last imputed dataset shown.

**Table 3:** Final recalibrated multivariable logistic regression model for predicting ECV ≥27%.

| Variable | Log odds | 95% CI | Odds ratio | Z statistic | P value |
| --- | --- | --- | --- | --- | --- |
| (Intercept) | -0.96 | -1.98 – 0.05 | 0.38 | -3.84 | <0.001 |
| BMI | -0.06 | -0.08 – -0.04 | 0.94 | -6.76 | <0.001 |
| eGFR | 0.01 | 0.01 – 0.02 | 1.01 | 3.45 | <0.001 |
| Female | 0.93 | 0.72 – 1.13 | 2.53 | 9.09 | <0.001 |
| ***ln* NT-proBNP** | **0.44** | **0.36 – 0.52** | **1.55** | **11.22** | **<0.001** |
| Stroke | -0.53 | -0.95 – -0.12 | 0.59 | -2.57 | 0.01 |
| White race | -0.45 | -0.74 – -0.17 | 0.64 | -3.22 | <0.01 |
| COPD | 0.41 | -0.01 – 0.83 | 1.50 | 1.94 | 0.05 |
| Prior HF hospitalisation | 0.29 | -0.04 – 0.61 | 1.33 | 1.78 | 0.07 |
| HTN | 0.19 | -0.02 – 0.41 | 1.21 | 1.79 | 0.07 |

ECV=extracellular volume, BMI=body mass index, CI=confidence interval, HTN= hypertension, COPD=chronic obstructive pulmonary disease, HF=heart failure, eGFR=estimated glomerular filtration rate, *ln*=natural logarithm, NT-proBNP=N-terminal pro B-type natriuretic peptide

### Prognostic value of ECV

Despite imperfect correlations between predicted vs. measured ECV (Figures 1 and 2), predicted ECV clearly identified vulnerable individuals at higher risk of adverse outcomes. Predicted ECV (continuous variable) associated with incident outcomes in all subgroups except for the smallest subgroup of the 174 patients with COPD (Supplemental Table 8). The Kaplan Meier plot for predicted ECV ≥27% or <27% in the combined derivation and validation cohorts are shown in Figure 3 where 483 adverse events (hospitalization for heart failure or death) occurred over a median of 1370 days. The univariable associations in Cox regression models between predicted ECV as a continuous variable and ECV ≥27% as a categorical variable are shown in Table 4. The hazard ratio and 95% CI for a 1 standard deviation (3.80 %) increase in ECV was 2.90 (2.45 – 3.44) and for ECV ≥27% was 2.21 (1.84 – 2.66).

**Figure 3:**
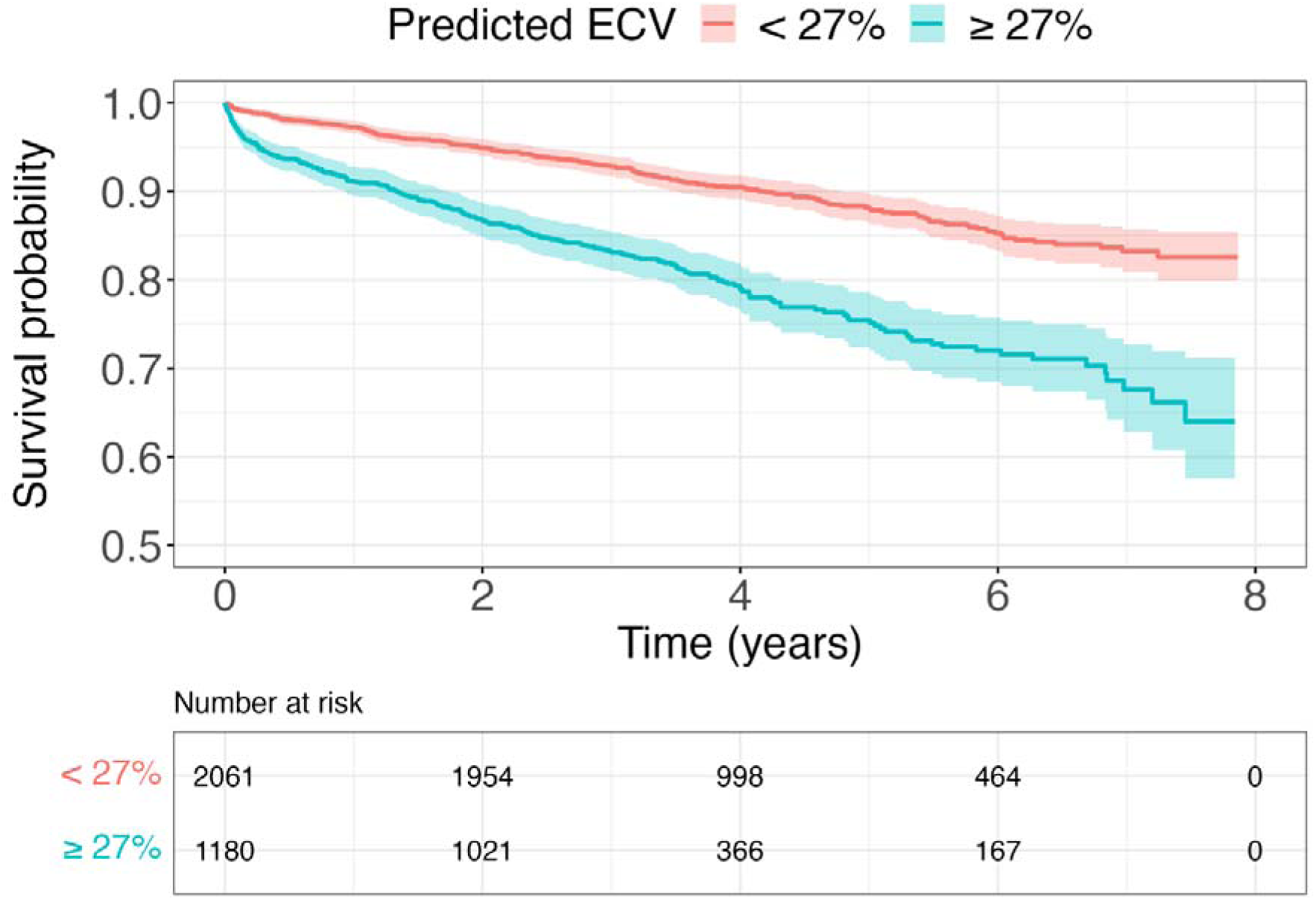
Kaplan Meier plots for predicted ECV ≥27% vs. <27% in 3241 individuals where 483 events occurred (i.e., hospitalization for heart failure or death) over a median follow-up of 1370 days.

**Table 4:** Univariable relationships between predicted ECV and composite outcome of heart failure hospitalisation and all-cause mortality in combined derivation and validation cohorts.

|  | **Hazard Ratio** | **95% CI** | **Chi square statistic** | **P value** |
| --- | --- | --- | --- | --- |
| Predicted ECV $\geq 27\%$ | 2.21 | 1.84 – 2.66 | 70.39 | <0.001 |
| Predicted ECV* | 2.90 | 2.45 – 3.44 | 150.06 | <0.001 |

\*Hazard ratio per 1 standard deviation (3.80 %) increase in ECV
CI=confidence interval

### Sample size estimates

Sample size estimates for the three different screening strategies are presented in Table 5 using: predicted ECV, measured ECV or no ECV, illustrating the numbers of individuals needed for screening and enrolment. These numbers follow the power calculations indicating that 1812 participants with an actual ECV ≥27% would be required to be enrolled in the hypothetical therapeutic trial (906 participants in each arm).

**Table 5:** Number of participants for a hypothetical clinical trial with an inclusion criterion of ECV ≥27%, whether predicted or measured with CMR with contrast. Three different screening strategies summarised. For the multivariable logistic regression model, sample size estimates for a probability threshold of 0.69 and 80% specificity are shown.

|  | **Screen with predicted ECV** | **Screen with CMR** | **No prior screening** |
| --- | --- | --- | --- |
| Screened | 6992 | 3940 | NA |
| Screened participants requiring CMR (%) | 0 (0 %) | 3940 (100%) | 0 (0%) |
| Enrolled | 2517 | 1812 | 3940 |
| Participants in trial with actual ECV $\geq 27\%$ (%) | 1812 (72%) | 1812 (100%) | 1812 (46%) |

Screening participants with the multivariable logistic regression predicted ECV model would obviate the need to perform CMR imaging prior to enrolment and thereby simplify screening for phase 3 trials. The model was able to predict ECV ≥27% with 70%, 80% and 90% specificity using various probability thresholds of 0.63, 0.69 and 0.78, respectively (Supplementary Figure 7 and Supplementary Tables 9 – 11). The specificity of the model can be selected depending on the clinical scenario. The varying sample size estimates for screening and enrolment using these specificity and probability thresholds are shown in Supplementary Table 12.

Using the model with 80% specificity as an illustrative example, 2517 participants with predicted ECV ≥27% would need to be enrolled to achieve the desired 1812 participants with an observed ECV ≥27% (Number of enrolled participants with predicted ECV ≥27% = number of enrolled participants with observed ECV ≥27% / positive predictive value, N= 1812/0.72). The prevalence of predicted ECV≥27% in the combined cohort was 36% and therefore 6992 patients would need to be screened to identify 2517 participants with predicted ECV ≥27% (Number of screened participants = number of enrolled participants with predicted ECV ≥27% / prevalence of predicted ECV ≥27%, N=2517/0.36).

Alternatively, all participants could be screened with a CMR scan prior to enrolment. The prevalence of observed ECV ≥27% was 46% in the combined cohorts. Therefore 3940 participants would need to be screened to identify the 1812 participants for enrolment. At the cost of 3940 additional scans, a CMR screening strategy would reduce the number of participants screened and enrolled by 3052 (43.6 % reduction) and 705 participants (28 % reduction), respectively, compared to screening with the model (Table 5).

Participants could be enrolled without prior screening. The prevalence of observed ECV ≥27% was 46% in the combined cohort, hence 3940 participants would need to be enrolled to achieve 1812 participants with actual ECV ≥27%. Opting out of screening would necessitate the enrolment of an additional 1423 (57 % increase) participants compared to screening with the model (Table 5).

## Discussion

In this work, we demonstrate that a variety of commonly available clinical covariates can predict myocardial ECV measures of MF without tomographic imaging such as CMR or CCT. Predicted ECV associated with ECV better than any other clinical variable. Predicted ECV as a continuous variable exhibited reasonable discrimination and calibration with external validation. Importantly, both predicted ECV and predicted ECV ≥27% (binary variable) associated with incident adverse events such as hospitalization for heart failure and death, confirming predicted ECV’s ability to identify vulnerable patients who may benefit from treatment. Finally, we provide sample size estimates for screening and enrolment to hypothetical phase 3 clinical trials testing an anti-fibrotic therapy using predicted ECV ≥27%, measured ECV ≥27% or no ECV. Supplemental data reveals the effect of choosing different specificities for predicted ECV ≥27% on these sample size estimates.

We believe predicted ECV represents the most efficient methodology to identify patients who may benefit from a proposed anti-fibrotic treatment compared to other clinical variables in our dataset. Efficient identification of those patients with elevated ECV who are likely to benefit from treatment is a crucial issue for pharmaceutical companies developing anti-fibrotic therapies. ECV ≥27% could represent a key eligibility criterion for anti-fibrotic treatment, enabling selective treatment of those likely to have MF rather than indiscriminate reliance on surrogate disease categories that identify patient with MF less robustly. Indeed, predicted ECV (acknowledging it is a composite measure) associated with measured ECV far better than any other clinical variable. ECV ≥27% was prevalent at just under 50%, whether measured or predicted, both in the entire cohort as well as various disease states (data not shown). Elevated ECV was not unique to any disease category. As such, these combined data suggest that there is a large proportion of patients that could benefit from efficacious anti-fibrotic treatment, not unique to any particular comorbidity. The threshold of ECV ≥27% represents the key eligibility criterion in the PIROUETTE Trial (13), where pirfenidone treatment lowered ECV and also NT-proBNP. In our cohorts, this threshold of ECV ≥27% identified vulnerable patients clearly at risk for hospitalization for heart failure or death.

These data may foster the design of a phase 3 clinical trial testing pharmacologic interventions targeting MF to demonstrate improved outcomes with treatment (e.g., reduced hospitalizations for heart failure and death). Promising therapeutics are already under various stages of development in phase 1 and phase 2 trial pipelines. Yet, in contrast to phase 2 trials (13), phase 3 trials cannot rely on advanced cardiac imaging to establish eligibility. A phase 3 trial without tomographic imaging involves certain trade-offs, with higher numbers of screened (77% increase) and enrolled (39% increase) participants, but without the complexities, costs, and risks of tomographic imaging that requires intravenous contrast exposure to measure ECV. While quantifying cost savings is beyond the scope of this manuscript (and will likely vary by region), we suspect the use of predicted ECV and the exclusion of ECV measured by tomographic imaging would yield considerable cost savings for a phase 3 trial, despite the requirement for higher numbers of screened and enrolled individuals. We reason that enrolment costs far exceed screening costs. Regardless, treatment eligibility cannot depend on tomographic imaging.

Natriuretic peptides and clinical covariates required for predicted ECV are readily available, easily measured, and a fraction of the cost of tomographic imaging that requires costly equipment, experienced specialty/referral centres, additional post processing, and clinical expertise. Furthermore, eligibility criteria for a proposed treatment would be simplified by excluding tomographic imaging. This is a critically important issue that would incentivize industry and justify the significant investment of resources required to develop anti-fibrotic treatments.

We acknowledge several limitations. First, ECV is an imperfect surrogate marker for fibrosis. The extracellular space can expand from other disease states such as cellular infiltration (e.g., from acute myocarditis (18) or amyloidosis(19)). We attempted to exclude these patients. We did not explore exclusion of amyloidosis specifically without tomographic imaging in this work which requires further study. Second, desired specificity thresholds for predicted ECV may vary in phase 3 trials as a function of costs and risks associated with the proposed treatment. We provided a range of specificities, but not an exhaustive list given space constraints. Estimated number of individuals required for screening and enrolment for a phase 3 trial depend on underlying assumptions. We attempted to remain conservative and employ reasonable assumptions regarding power, effect size, event rates, and other parameters similar to a recent phase 3 trial.(17) Finally, we believed formal cost effectiveness analysis was beyond the scope of this manuscript.

## Conclusions

Ubiquitous and inexpensive data elements, such as comorbidity and natriuretic peptides, permit prediction of myocardial ECV with reasonable discrimination and calibration in external validation cohorts. Predicted ECV ≥27% was prevalent and associated with adverse outcomes. Predicted ECV ≥27% permits efficient identification of vulnerable patients who may benefit from selective treatment rather than indiscriminate reliance on surrogate disease categories. Predicted ECV may foster the design of phase 3 trials of anti-fibrotic therapies, without the costs, complexity and risk of tomographic imaging, but with higher numbers of screened and enrolled participants.

## Data Availability

All data produced in the present study are available upon reasonable request to the authors

