## Supplemental material for "Development and validation of imaging-free myocardial fibrosis prediction models, association with outcomes, and sample size estimation for phase 3 trials"

**Supplementary Table 1:** Table of missing values

|  | **Derivation**  **(N = 1663)** | **Validation**  **(N = 1578)** | **Overall**  **(N = 3241)** |
| --- | --- | --- | --- |
| **Demographics** |  |  |  |
| Age (%) | 0 (0.0) | 0 (0.0) | 0 (0.0) |
| Female (%) | 0 (0.0) | 0 (0.0) | 0 (0.0) |
| White race (%) | 0 (0.0) | 0 (0.0) | 0 (0.0) |
| Body mass index (%) | 16 (1.0) | 0 (0.0) | 16 (0.5) |
| HF stage (%) | 0 (0.0) | 0 (0.0) | 0 (0.0) |
| **Comorbidity** |  |  |  |
| Prior heart failure admission (%) | 0 (0.0) | 0 (0.0) | 0 (0.0) |
| MI (%) | 0 (0.0) | 401 (25.4) | 401 (12.4) |
| Stroke or TIA (%) | 0 (0.0) | 0 (0.0) | 0 (0.0) |
| Diabetes (%) | 0 (0.0) | 0 (0.0) | 0 (0.0) |
| Hypertension (%) | 0 (0.0) | 0 (0.0) | 0 (0.0) |
| Hyperlipidaemia (%) | 0 (0.0) | 0 (0.0) | 0 (0.0) |
| Atrial fibrillation (%) | 0 (0.0) | 0 (0.0) | 0 (0.0) |
| COPD (%) | 0 (0.0) | 0 (0.0) | 0 (0.0) |
| Current smoker (%) | 0 (0.0) | 0 (0.0) | 0 (0.0) |
| Ex-smoker (%) | 0 (0.0) | 0 (0.0) | 0 (0.0) |
| QRS duration (%) | 349 (21.0) | 1078 (68.3) | 1427 (44.0) |
| **Laboratory measurements** |  |  |  |
| eGFR (%) | 22 (1.3) | 0 (0.0) | 22 (0.7) |
| *ln*NT-proBNP (%) | 413 (24.8) | 181 (11.5) | 594 (18.3) |
| **CMR characteristics** |  |  |  |
| Left ventricle ejection fraction (%) | 3 (0.1) | 0 (0.0) | 3 (0.1) |
| Left ventricle mass index (%) | 10 (0.6) | 0 (0.0) | 10 (0.3) |
| Global longitudinal strain (%) | 58 (3.5) | 0 (0.0) | 58 (1.8) |
| Infarct LGE (%) | 0 (0.0) | 0 (0.0) | 0 (0.0) |
| Myocardial ECV percentage (%) | 237 (14.3) | 0 (0.0) | 237 (7.3) |

Missing values are n (%).

HF=heart failure, MI=myocardial infarction, TIA=transient ischaemic attack, COPD=chronic obstructive pulmonary disease, eGFR=estimated glomerular filtration rate, NT-proBNP=N-terminal pro B-type natriuretic peptide, LGE=late gadolinium enhancement, ECV=extracellular volume

**Supplementary Figure 1:** Model assumptions for multivariable linear regression model for predicting ECV. Illustrative data from last imputed dataset.

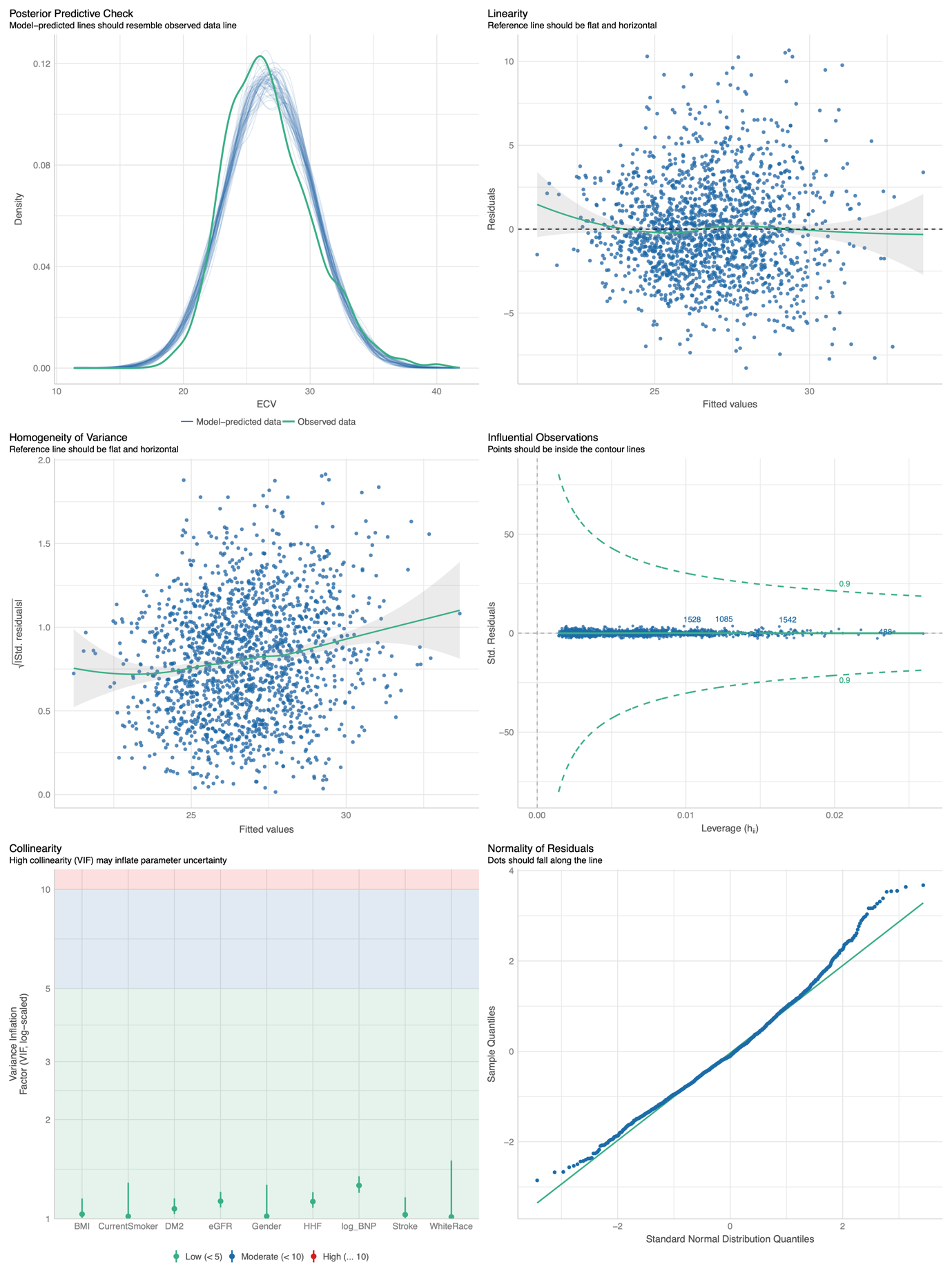

BMI

Current

smoker

DM

eGFR

Female sex

Prior HF hospitalisation

Ln

NT-proBNP

Stroke

White

race

**Supplementary Figure 2:** Model assumptions for multivariable logistic regression model for predicting ECV. Illustrative data from last imputed dataset.

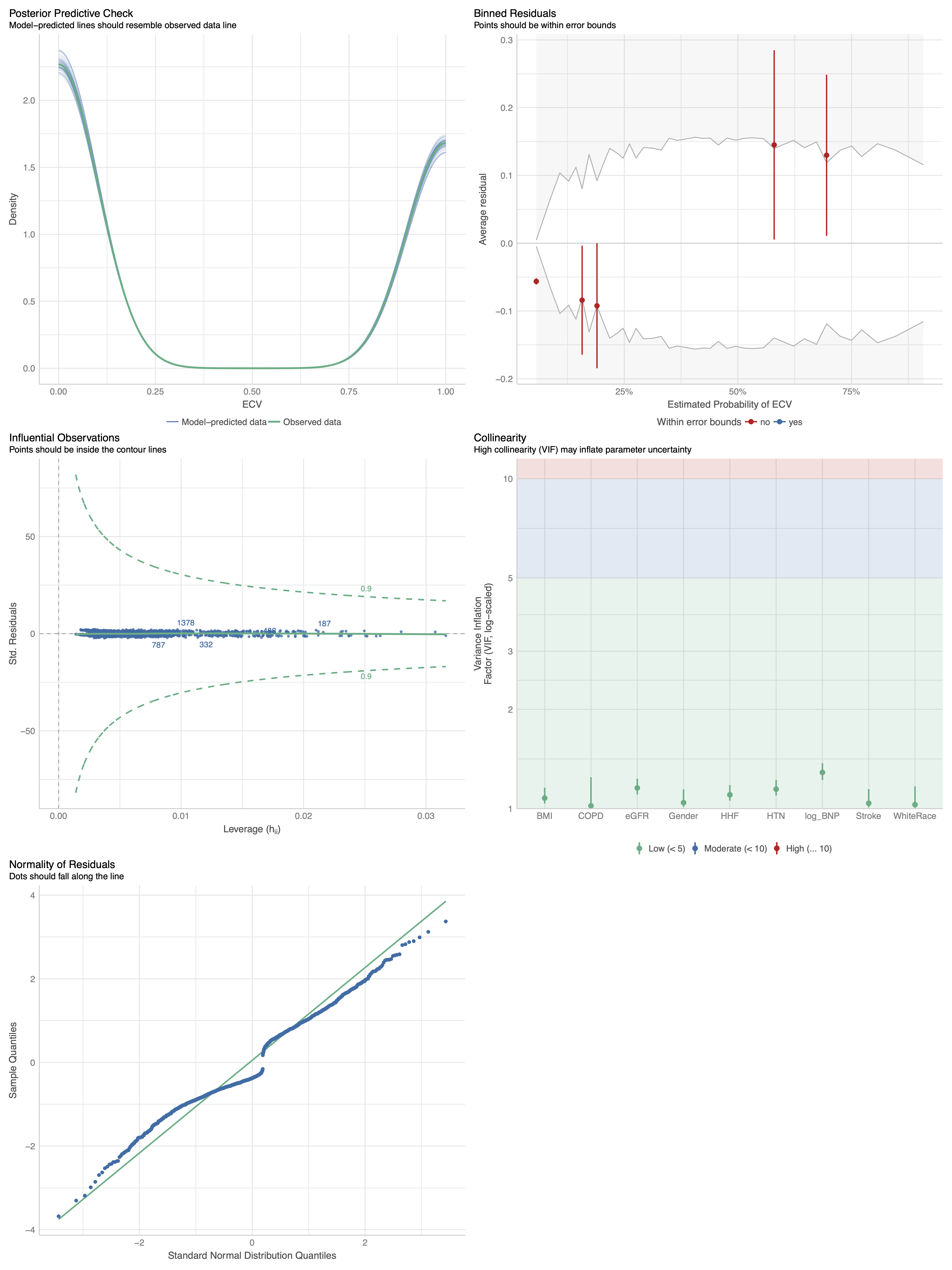

BMI

COPD

eGFR

Female sex

Prior HF hospitalisation

Ln

NT-proBNP

Stroke

White

race

HTN

**Supplementary Table 2:** Univariable associations between ECV as a continuous variable. B-type natriuretic peptide exhibited the strongest association among solitary predictor variables.

| **Variable** | **Intercept** | **Regression coefficient (standard error)** | **95% confidence interval** | **T statistic** | **P value** |
| --- | --- | --- | --- | --- | --- |
| Age | 25.48 | 0.02 (0.01) | 0.01– 0.03 | 4.77 | <0.001 |
| Female | 26.04 | 1.88 (0.17) | 1.54 – 2.22 | 10.79 | <0.001 |
| White race | 27.00 | -0.26 (0.25) | -0.75 – 0.22 | -1.07 | 0.28 |
| BMI | 30.11 | -0.12 (0.01) | -0.15 – -0.09 | -8.00 | <0.001 |
| MI | 26.63 | 0.83 (0.23) | 0.37 – 1.29 | 3.53 | <0.001 |
| Stroke | 26.77 | 0.09 (0.36) | -0.62 – 0.80 | 0.24 | 0.81 |
| DM | 26.70 | 0.67 (0.29) | 0.10 – 1.25 | 2.30 | 0.02 |
| HTN | 26.71 | 0.18 (0.18) | -0.18 – 0.54 | 0.96 | 0.34 |
| Hyperlipidaemia | 26.68 | 0.29 (0.19) | -0.08 – 0.66 | 1.55 | 0.12 |
| COPD | 26.71 | 1.17 (0.37) | 0.44 – 1.90 | 3.13 | <0.01 |
| AF | 26.69 | 0.63 (0.26) | 0.12 – 1.14 | 2.42 | 0.02 |
| Current smoker | 26.65 | 1.08 (0.28) | 0.52 – 1.64 | 3.79 | <0.001 |
| Ex-smoker | 26.75 | 0.07 (0.19) | -0.30 – 0.44 | 0.39 | 0.69 |
| Prior HF hospitalisation | 26.57 | 1.79 (0.28) | 1.24 – 2.33 | 6.42 | <0.001 |
| eGFR | 27.45 | -0.01 (0.01) | -0.02 – 0.00 | -1.26 | 0.21 |
| ***ln* NT-proBNP** | **22.42** | **0.86 (0.05)** | **0.76 – 0.97** | **16.22** | **<0.001** |
| QRS duration | 26.10 | 0.01 (0.00) | 0.00 – 0.01 | 1.61 | 0.11 |
| **Predicted ECV** | **-0.72** | **0.96 (0.04)** | **0.88 – 1.03** | **23.83** | **<0.001** |

ECV=extracellular volume, BMI=body mass index, MI=myocardial infarction, DM= diabetes mellitus, HTN= hypertension, COPD=chronic obstructive pulmonary disease, AF=atrial fibrillation, HF=heart failure, eGFR=estimated glomerular filtration rate, ln=natural logarithm, NT-proBNP=N-terminal pro B-type natriuretic peptide

**Supplementary Table 3:** Regression coefficients for multivariable linear model for predicting ECV as a continuous variable

|  | **Original model (95% CI)** | **Optimism adjusted model (95% CI)** | **Recalibrated model (95% CI)** |
| --- | --- | --- | --- |
| Intercept | 23.26 (21.73 – 24.79) | 23.02 (21.51 ­– 24.53) | 25.09 (23.47 – 26.70) |
| BMI | -0.11 (-0.14 – -0.08) | -0.11 (-0.14 – -0.08) | -0.12 (-0.14 – -0.09) |
| Current smoker | 0.78 (0.29 – 1.28) | 0.77 (0.28 – 1.27) | 0.82 (0.29 – 1.34) |
| eGFR | 0.02 (0.01 – 0.04) | 0.02 (0.01 – 0.04) | 0.03 (0.01 – 0.04) |
| Female | 1.81 (1.50 – 2.12) | 1.79 (1.49 – 2.10) | 1.90 (1.57 – 2.22) |
| Prior HF hospitalisation | 0.60 (0.09 – 1.11) | 0.59 (0.09 – 1.10) | 0.63 (0.09 – 1.17) |
| *ln* NT-proBNP | 0.87 (0.76 – 0.99) | 0.86 (0.75 – 0.98) | 0.91 (0.80 – 1.03) |
| White race | -0.74 (-1.16 – -0.31) | -0.73 (-1.15 – -0.31) | -0.77 (-1.22 – -0.32) |
| Stroke | -0.55 (-1.18 – 0.08) | -0.54 (-1.17 – 0.08) | -0.57 (-1.24 – 0.09) |
| DM | 0.58 (0.06 – 1.09) | 0.57 (0.06 – 1.08) | 0.60 (0.06 – 1.15) |

ECV=extracellular volume, BMI=body mass index, CI=confidence interval, DM= diabetes mellitus, HF=heart failure, eGFR=estimated glomerular filtration rate, ln=natural logarithm, NT-proBNP=N-terminal pro B-type natriuretic peptide

**Supplementary Table 4:** Validation metrics of multivariable linear model for predicting ECV

|  | **Internal validation** | | **External validation** | | **Recalibration** | |
| --- | --- | --- | --- | --- | --- | --- |
|  | **Estimate (SE)** | **95% CI** | **Estimate (SE)** | **95% CI** | **Estimate (SE)** | **95% CI** |
| **R square** | 0.29 (0.02) | 0.26 – 0.33 | 0.24 (0.02) | 0.20 – 0.28 | 0.24 (0.02) | 0.20 – 0.28 |
| **Calibration slope** | 0.99 (0.003) | 0.98 – 1.0 | 1.06 (0.05) | 0.96 – 1.16 | 1.00 (0.05) | 0.91 – 1.10 |
| **Calibration intercept** | 0.28 (0.08) | 0.14 – 0.44 | 0.76 (1.32) | -1.85 – 3.33 | -0.38 (1.36) | -2.67 – 2.65 |
| **Mean absolute error** | 2.24 (0.04) | 2.16 – 2.33 | 3.12 (0.07) | 2.99 – 3.26 | 2.66 (0.06) | 2.55 – 2.78 |
| **Root mean square error** | 2.87 (0.06) | 2.76 – 2.98 | 4.14 (0.10) | 3.94 – 4.34 | 3.48 (0.09) | 3.32 – 3.66 |

CI=confidence interval, SE=standard error

**Supplementary Figure 3:** Calibration plot of original multivariable linear model for predicting ECV in derivation cohort. Plot from last imputed dataset shown.

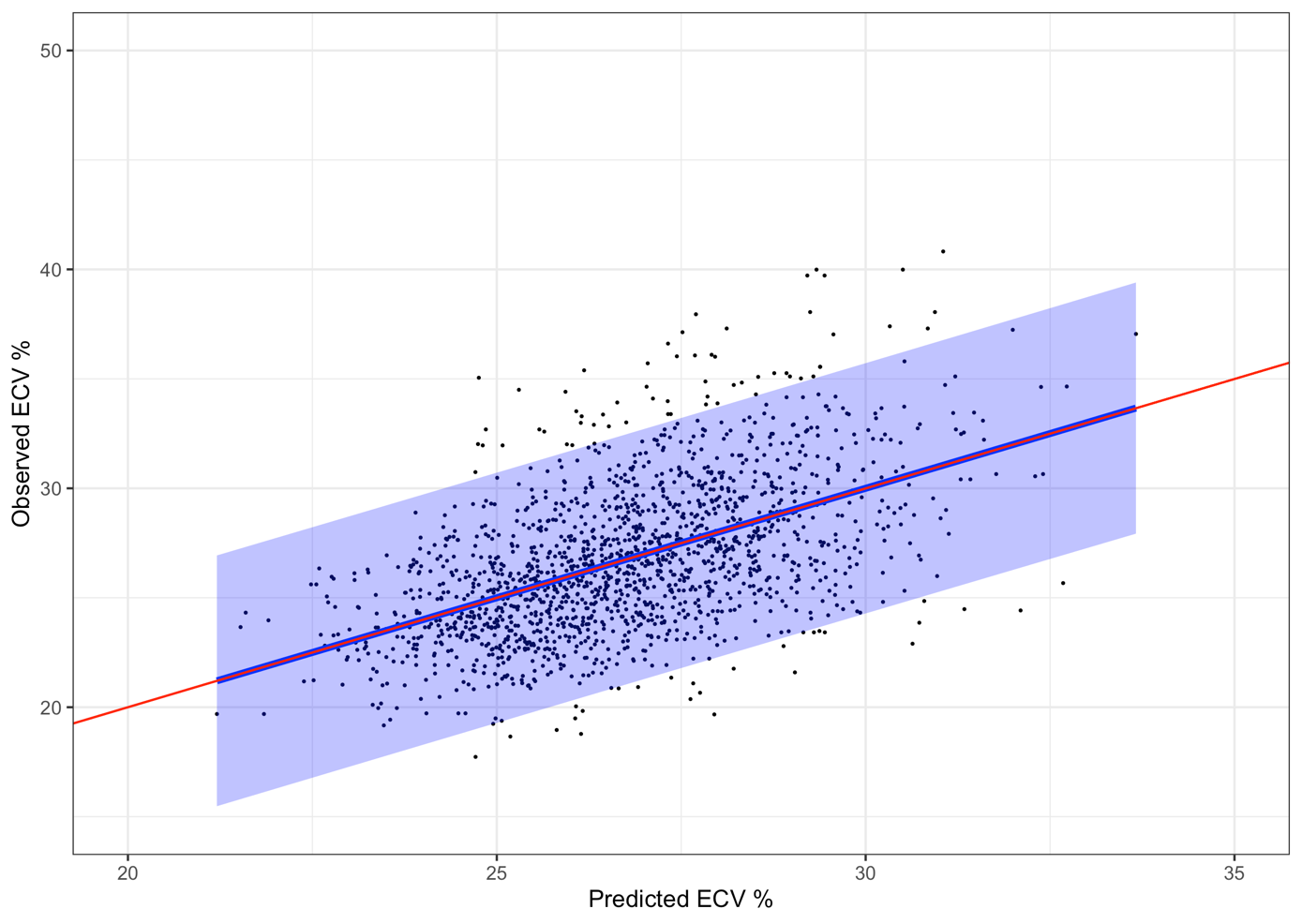

**Supplementary Figure 4:** Calibration plot of optimism adjusted multivariable linear model for predicting ECV in validation cohort. Plot from last imputed dataset shown.

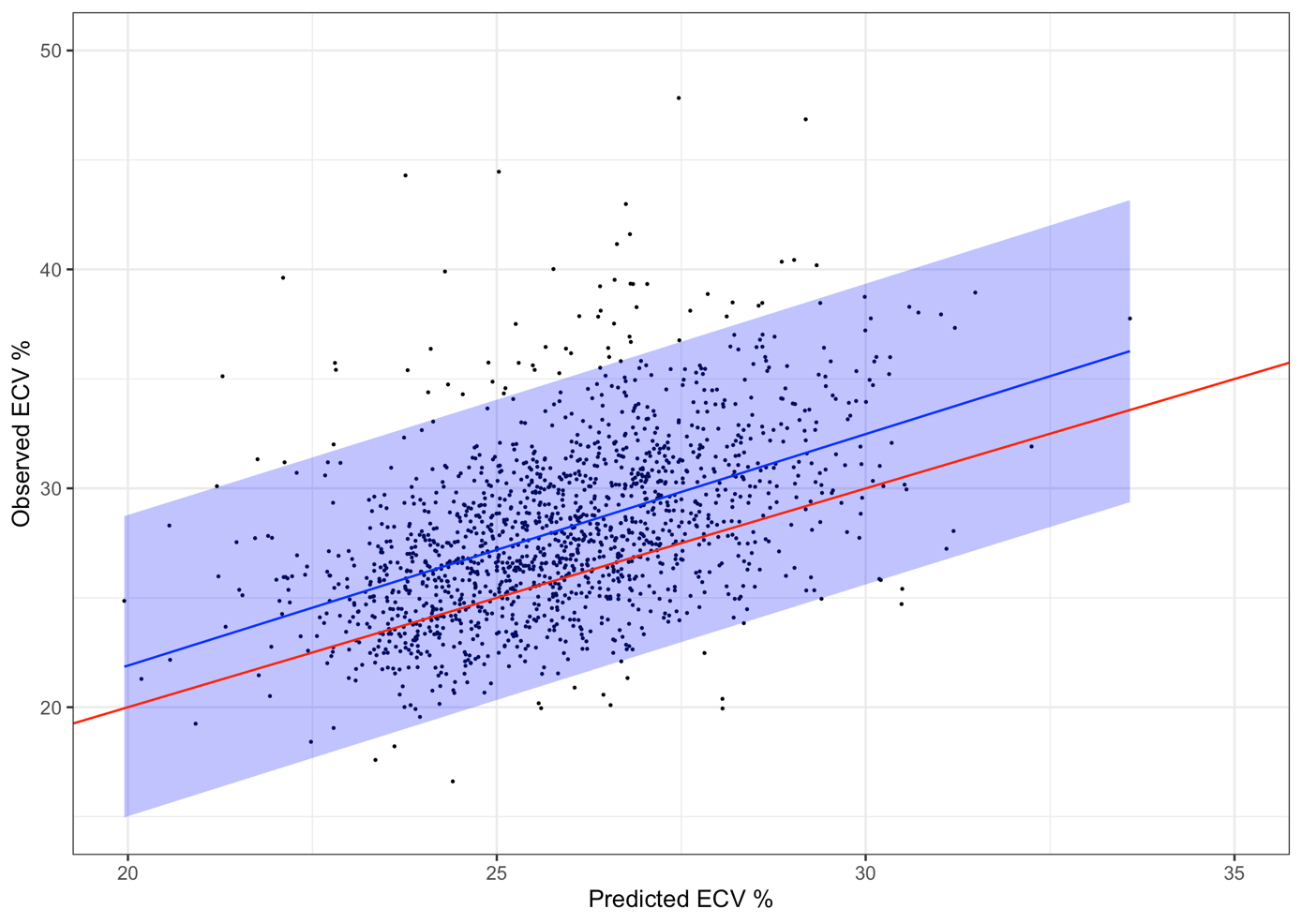

**Supplementary Table 5** Univariable associations between candidate predictor variables and ECV ≥ 27%.

| **Variable** | **Intercept** | **Log odds (SE)** | **95% CI** | **Odds ratio** | **Z statistic** | **p value** |
| --- | --- | --- | --- | --- | --- | --- |
| Age | -1.02 | 0.01 (0.003) | 0.01 – 0.02 | 1.01 | 4.36 | <0.001 |
| Female | -0.70 | 0.98 (0.11) | 0.76 – 1.20 | 2.67 | 8.88 | <0.001 |
| White Race | -0.16 | -0.17 (0.15) | -0.46 – 0.12 | 0.85 | -1.14 | 0.26 |
| BMI | 1.51 | -0.06 (0.01) | -0.08 – -0.05 | 0.94 | -6.48 | <0.001 |
| MI | -0.38 | 0.42 (0.14) | 0.15 – 0.70 | 1.53 | 3.05 | <0.01 |
| Stroke | -0.29 | -0.22 (0.22) | -0.65 – 0.21 | 0.80 | -1.01 | 0.31 |
| DM | -0.32 | 0.15 (0.17) | -0.19 – 0.49 | 1.16 | 0.87 | 0.38 |
| HTN | -0.38 | 0.21 (0.11) | -0.01 – 0.42 | 1.23 | 1.91 | 0.06 |
| Hyperlipidaemia | -0.37 | 0.20 (0.11) | -0.01 – 0.42 | 1.23 | 1.88 | 0.06 |
| COPD | -0.34 | 0.66 (0.22) | 0.23 – 1.09 | 1.93 | 3.00 | <0.01 |
| AF | -0.35 | 0.37 (0.16) | 0.07 – 0.68 | 1.45 | 2.41 | 0.02 |
| Current smoker | -0.34 | 0.29 (0.16) | -0.03 – 0.61 | 1.34 | 1.78 | 0.08 |
| Ex smoker | -0.34 | 0.11 (0.11) | -0.11 – 0.32 | 1.11 | 0.96 | 0.34 |
| Prior HF hospitalisation | -0.40 | 0.85 (0.17) | 0.53 – 1.18 | 2.35 | 5.13 | <0.001 |
| eGFR | 0.00 | -0.004 (0.004) | -0.01 – 0.00 | 1.00 | -0.96 | 0.34 |
| ***ln* NT-proBNP** | **-2.69** | **0.47 (0.04)** | **0.39 – 0.55** | **1.60** | **11.69** | **<0.001** |
| QRS duration | -0.45 | 0.001 (0.002) | 0.00 – 0.01 | 1.00 | 0.62 | 0.54 |
| **Predicted ECV** | **-17.51** | **0.59 (0.04)** | **0.52 – 0.67** | **1.81** | **15.53** | **<0.001** |
| **Predicted ECV ≥ 27%*** | **-1.13** | **1.80 (0.12)** | **1.56 – 2.04** | **6.05** | **14.77** | **<0.001** |

*Multivariable logistic regression model with 80% specificity

ECV=extracellular volume, BMI=body mass index, CI=confidence interval, MI=myocardial infarction, DM= diabetes mellitus, HTN= hypertension, COPD=chronic obstructive pulmonary disease, AF=atrial fibrillation, HF=heart failure, eGFR=estimated glomerular filtration rate, ln=natural logarithm, NT-proBNP=N-terminal pro B-type natriuretic peptide, SE=standard error

**Supplementary Table 6:** Regression coefficients for multivariable logistic regression model for predicting ECV ≥27%

|  | **Original model (95% CI)** | **Optimism adjusted model (95% CI)** | **Recalibrated model (95% CI)** |
| --- | --- | --- | --- |
| (Intercept) | -2.40 (-3.63 – -1.17) | -2.34 (-3.53 – -1.15) | -0.96 (-1.98 – 0.05) |
| BMI | -0.08 (-0.10 – -0.05) | -0.08 (-0.10 – -0.05) | -0.06 (-0.08 – -0.04) |
| eGFR | 0.02 (0.01 – 0.03) | 0.02 (0.01 – 0.03) | 0.01 (0.01 – 0.02) |
| Female | 1.15 (0.90 – 1.40) | 1.12 (0.88 – 1.36) | 0.93 (0.72 – 1.13) |
| *ln* NT-proBNP | 0.54 (0.45 – 0.64) | 0.53 (0.44 – 0.62) | 0.44 (0.36 – 0.52) |
| Stroke | -0.66 (-1.16 – -0.16) | -0.64 (-1.13 – -0.15) | -0.53 (-0.95 – -0.12) |
| White race | -0.56 (-0.91 – -0.22) | -0.55 (-0.88 – -0.21) | -0.45 (-0.74 – -0.17) |
| COPD | 0.50 (-0.01 – 1.01) | 0.49 (0.00 – 0.99) | 0.41 (-0.01 – 0.83) |
| Prior HF hospitalisation | 0.36 (-0.04 – 0.75) | 0.35 (-0.03 – 0.73) | 0.29 (-0.04 – 0.61) |
| HTN | 0.24 (-0.02 – 0.50) | 0.23 (-0.02 – 0.49) | 0.19 (-0.02 – 0.41) |

ECV=extracellular volume, BMI=body mass index, CI=confidence interval, HTN= hypertension, COPD=chronic obstructive pulmonary disease, HF=heart failure, eGFR=estimated glomerular filtration rate, ln=natural logarithm, NT-proBNP=N-terminal pro B-type natriuretic peptide

**Supplementary Table 7:** Validation metrics of multivariable logistic regression model for predicting ECV ≥27%

|  | **Internal validation** | | **External validation** | | **Recalibration** | |
| --- | --- | --- | --- | --- | --- | --- |
|  | **Estimate (SE)** | **95% CI** | **Estimate (SE)** | **95% CI** | **Estimate (SE)** | **95% CI** |
| **C-statistic** | 0.78 (0.01) | 0.75 – 0.80 | 0.74 (0.01) | 0.71 – 0.76 | 0.74 (0.01) | 0.71 – 0.76 |
| **Calibration slope** | 0.97 (0.01) | 0.96 – 0.99 | 0.83 (0.06) | 0.71 – 0.95 | 1.00 (0.07) | 0.85 – 1.15 |
| **Calibration intercept** | -0.01 (0.01) | -0.02 – 0.01 | 0.97 (0.08) | 0.82 – 1.13 | 0.00 (0.06) | -0.11 – 0.11 |

CI=confidence interval, SE=standard error

**Supplementary Figure 5:** Calibration plot of original multivariable logistic regression model for predicting ECV ≥27% in derivation cohort. Plot from last imputed dataset shown.

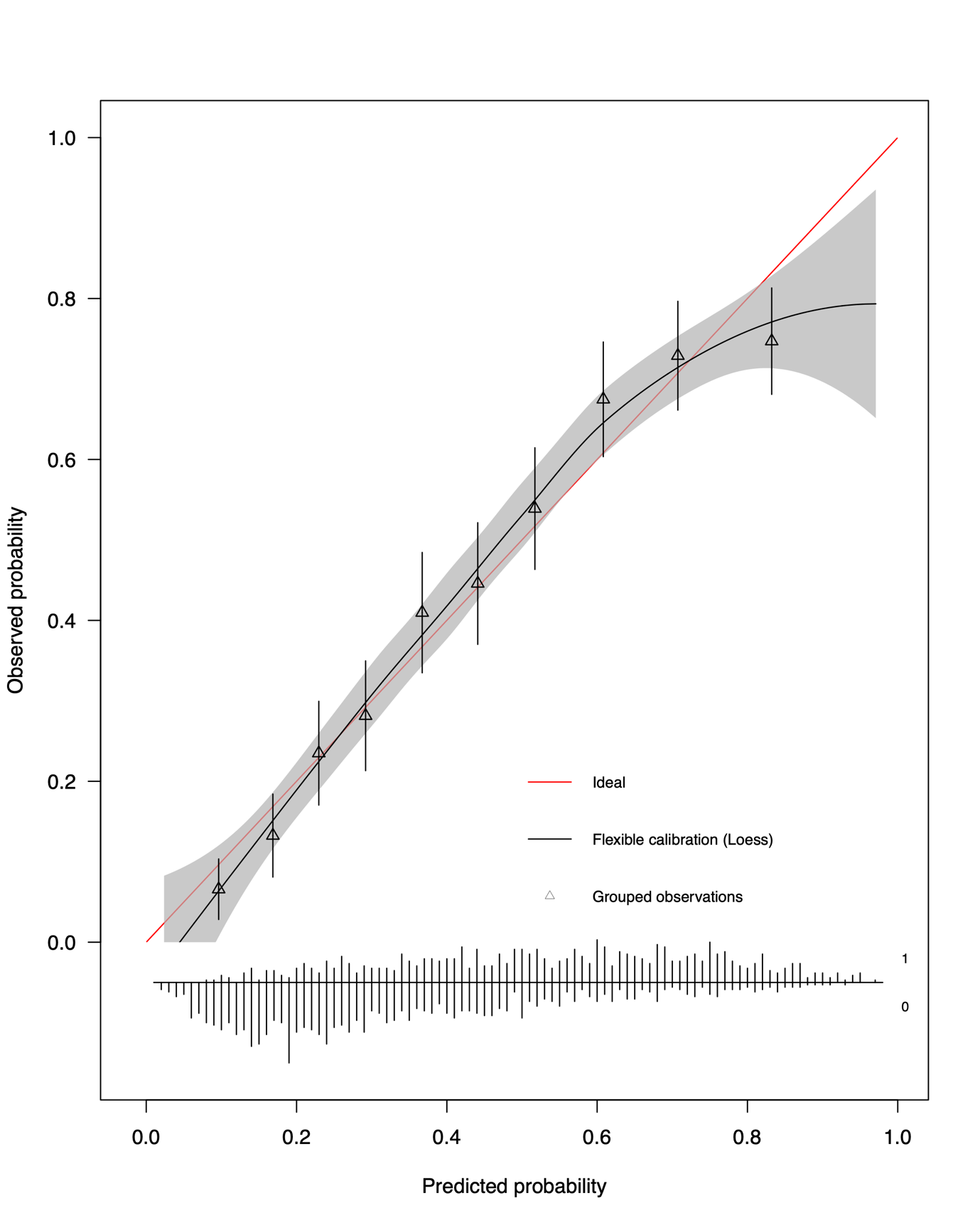

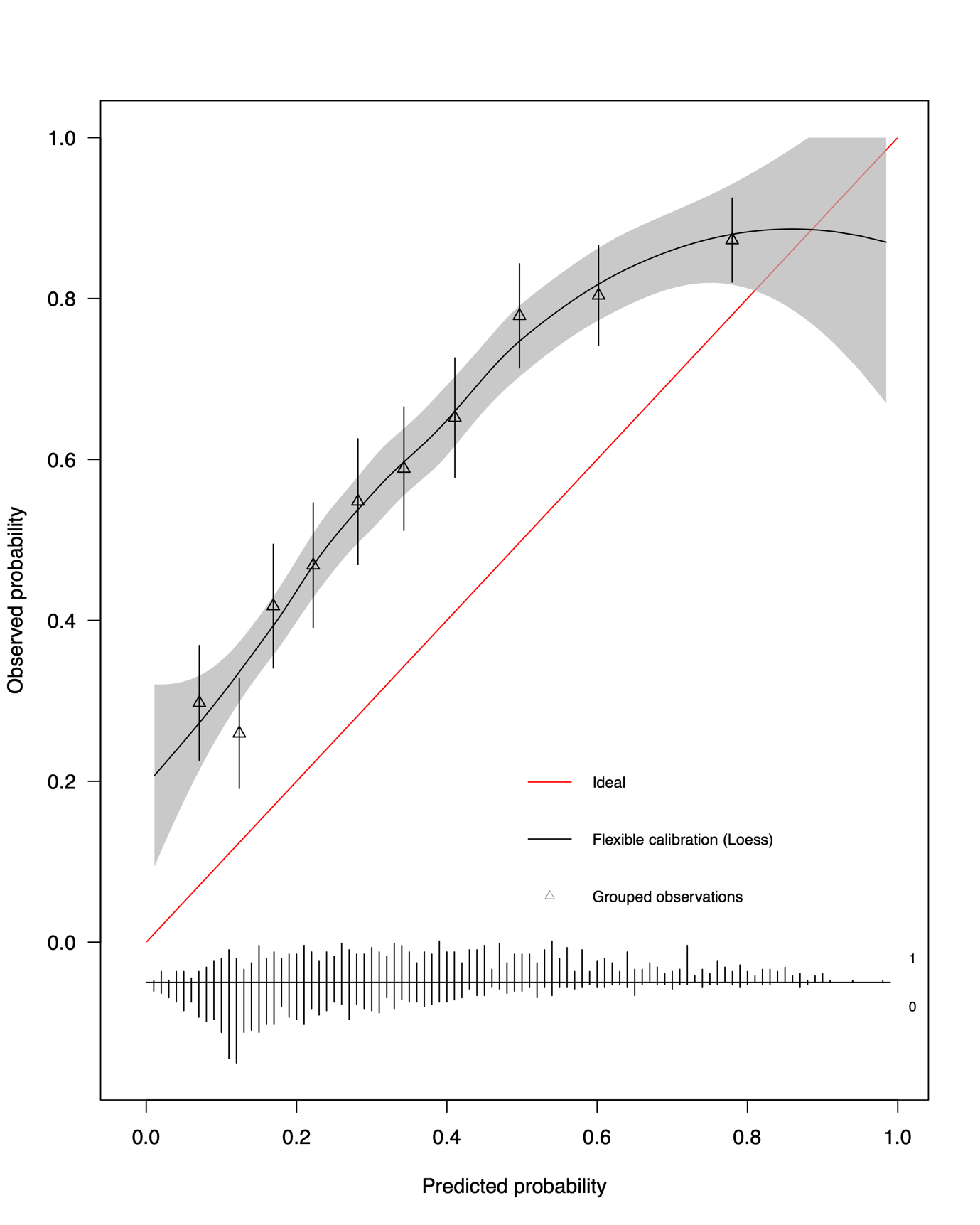
**Supplementary Figure 6:** Calibration plot of optimism corrected multivariable logistic regression model for predicting ECV ≥27% in validation cohort. Plot from last imputed dataset shown.

**Supplemental Table 8:** Predicted ECV, expressed as a binary variable (ECV ≥27%) or as a continuous variable, associates with incident outcomes in Cox regression models in various disease categories with the exception of the relatively small numbers of patients with COPD (n=174).

|  | **Measured ECV ≥ 27%** | | | **Predicted ECV ≥ 27%** | | | **Measured ECV*** | | | **Predicted ECV*** | | |
| --- | --- | --- | --- | --- | --- | --- | --- | --- | --- | --- | --- | --- |
|  | HR and 95% CI | Chi square  statistic | P value | HR and 95% CI | Chi square  statistic | P value | HR and 95% CI | Chi square  statistic | P value | HR and 95% CI | Chi square  statistic | P value |
| All patients (n=3241) | 2.63 (2.14 – 3.22) | 86.61 | <0.001 | 2.21 (1.84 – 2.66) | 70.39 | <0.001 | 1.70 (1.58 – 1.83) | 195.40 | <0.001 | 1.74 (1.59 – 1.90) | 150.06 | <0.001 |
| Diabetes (n=495) | 2.20 (1.51 – 3.19) | 17.26 | <0.001 | 1.77 (1.27 – 2.46) | 11.66 | <0.001 | 1.63 (1.40 – 1.90) | 40.48 | <0.001 | 1.45 (1.24 – 1.69) | 22.68 | <0.001 |
| Hypertension (n=1416) | 2.43 (1.88 – 3.12) | 47.19 | <0.001 | 2.01 (1.59 – 2.54) | 34.74 | <0.001 | 1.63 (1.48 – 1.80) | 96.40 | <0.001 | 1.61 (1.44 - 1.81) | 66.73 | <0.001 |
| HHF (n=354) | 1.47 (0.97 – 2.24) | 3.37 | 0.07 | 1.14 (0.79 – 1.64) | 0.47 | 0.494 | 1.39 (1.18 – 1.64) | 15.09 | <0.001 | 1.25 (1.06 – 1.49) | 6.77 | 0.01 |
| LGE MI (n=637) | 2.27 (1.64 – 3.13) | 25.19 | <0.001 | 1.89 (1.41 – 2.54) | 18.30 | <0.001 | 1.69 (1.48 – 1.94) | 56.78 | <0.001 | 1.62 (1.39 – 1.87) | 40.69 | <0.001 |
| AF (n=369) | 2.62 (1.63 – 4.22) | 16.19 | <0.001 | 2.26 (1.45 – 3.52) | 13.51 | <0.001 | 1.94 (1.58 – 2.38) | 40.48 | <0.001 | 1.60 (1.32 – 1.93) | 24.09 | <0.001 |
| COPD (n=174) | 2.45 (1.26 – 4.79) | 7.19 | 0.01 | 1.15 (0.67 – 1.97) | 0.25 | 0.62 | 1.59 (1.24 – 2.05) | 13.44 | <0.001 | 1.27 (1.00 – 1.62) | 3.86 | 0.06 |
| Stage A HF (n=862) | 1.82 (1.12 – 2.95) | 6.16 | 0.016 | 2.00 (1.18 – 3.39) | 6.85 | 0.011 | 1.42 (1.14 – 1.77) | 9.89 | 0.002 | 1.39 (1.07 – 1.82) | 6.16 | 0.016 |
| Stage B HF (n=855) | 2.06 (1.36 – 3.13) | 11.86 | <0.001 | 1.86 (1.23 – 2.81) | 8.98 | 0.003 | 1.68 (1.40 – 2.02) | 31.93 | <0.001 | 1.56 (1.27 – 1.91) | 18.38 | <0.001 |
| Stage C HF (n=1157) | 2.32 (1.74 – 3.07) | 34.09 | <0.001 | 1.53 (1.20 – 1.96) | 11.71 | <0.001 | 1.53 (1.39 – 1.68) | 74.26 | <0.001 | 1.46 (1.30 – 1.64) | 41.03 | <0.001 |

*HR per 1 standard deviation increase in measured ECV (3.80 %) or predicted ECV (1.98 %)

**Supplementary Figure 7:** Illustrative density plot from final imputed dataset. Red and blue curves shows density plots of patients with observed ECV <27% and ≥27%, respectively. The output of the multivariable logistic regression model i.e. probability of ECV ≥27%, is shown on the horizontal axis. The vertical lines represent probability thresholds of 0.63, 0.69 and 0.78, corresponding to specificities of categorising ECV ≥27 of 70%, 80% and 90% respectively.

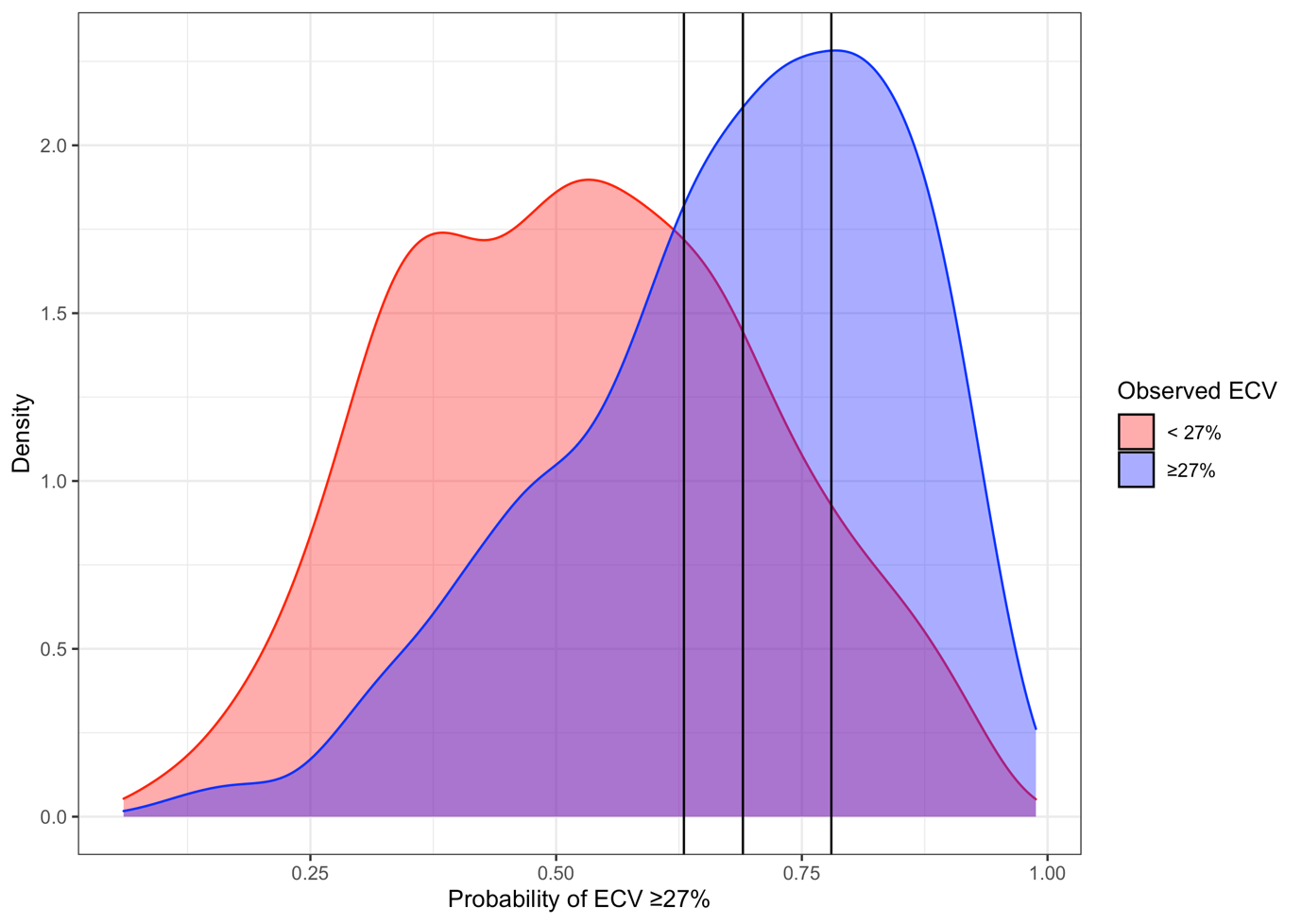

**Supplementary Table 9:** Illustrative table of observed vs. predicted ECV >27% from final imputed dataset using probability threshold of 0.63 as selection criteria for multivariable logistic regression model.

|  | **Predicted ECV <27%** | **Predicted ECV ≥27%** |
| --- | --- | --- |
| **Observed ECV < 27%** | 1139 | 491 |
| **Observed ECV ≥ 27%** | 559 | 1052 |

Specificity = 70%

Sensitivity = 65%

Positive predictive value = 68%

Negative predictive value = 67%

**Supplementary Table 10:** Illustrative table of observed vs. predicted ECV >27% from final imputed dataset using probability threshold of 0.69 as selection criteria for multivariable logistic regression model.

|  | **Predicted ECV <27%** | **Predicted ECV ≥27%** |
| --- | --- | --- |
| **Observed ECV < 27%** | 1304 | 326 |
| **Observed ECV ≥ 27%** | 757 | 854 |

Sensitivity = 53%

Specificity = 80%

Positive predictive value = 72%

Negative predictive value = 63%

**Supplementary Table 11:** Illustrative table of observed vs. predicted ECV >27% from final imputed dataset using probability threshold of 0.78 as selection criteria for multivariable logistic regression model.

|  | **Predicted ECV <27%** | **Predicted ECV ≥27%** |
| --- | --- | --- |
| **Observed ECV < 27%** | 1466 | 164 |
| **Observed ECV ≥ 27%** | 1069 | 542 |

Specificity = 90%

Sensitivity = 34%

Positive predictive value = 77%

Negative predictive value = 58%

**Supplementary Table 12:** Number of participants for a hypothetical clinical trial with an inclusion criterion of ECV ≥27%. Multivariable logistic regression models with probability thresholds of 0.63, 0.69 and 0.78 shown.

|  | **Probability threshold 0.63**  Specificity 70%  Sensitivity 65% | **Probability threshold 0.69**  Specificity 80%  Sensitivity 53% | **Probability threshold 0.78**  Specificity 90%  Sensitivity 34% |
| --- | --- | --- | --- |
| Screened | 5599 | 6992 | 10,848 |
| Enrolled | 2665 | 2517 | 2354 |
| Participants in trial with observed ECV ≥27% (%) | 1812 (68%) | 1812 (72%) | 1812 (77%) |
